# Genetic and Epidemiologic Assessment of Mandibular Cortical Indices and Bone Mineral Density in Peripubertal Children: The Generation R Study

**DOI:** 10.1101/2025.05.16.25327746

**Authors:** Vid Prijatelj, Olja Grgic-Chavez, Justin van der Tas, Constanza L. Andaur Navarro, Andre G. Uitterlinden, Fernando Rivadeneira, Eppo B. Wolvius, Carolina Medina-Gomez

## Abstract

**Objective:** The panoramic mandibular index (PMI) and mental index (MI) assessed on dental panoramic radiographs (DPRs) have been postulated as useful for the assessment of adult bone health. However, their utility in children remains to be determined. Our objective was to establish genetic determinants of the PMI/MI and to evaluate the relationship between these indices and total body less-head bone mineral density (TBLH-BMD) by leveraging data from medical records and genetic profiles of Dutch children.

**Study design:** This study was embedded in the Generation R Study including 3,518 participants at a mean age of 13 years. BMD was obtained from dual-energy X-ray (DXA) scans, while radiomorphometric measurements of the mandibular bone were obtained from DPRs. Genome-wide association studies (GWAS) on PMI/MI were performed using individual genotyped data imputed to the 1000 Genomes reference panel. The association between PMI/MI and BMD was comprehensively assessed following a combined observational and genetic analysis, both corrected for biological covariates such as sex, age, and others, using a BMD polygenic risk score (PGS) in pubescent children.

**Results:** The PMI and MI GWAS identified an associated signal (p=2.53×10^-9^) mapping to the *ODF3/BET1L/RIC8A/SIRT3* locus, previously associated with BMD. Moreover, significant differences in PMI and MI were observed across the extremes of the TBLH-BMD PGS distribution. Our results also show that a standard deviation (SD) increase in measured TBLH-BMD was associated with 0.244 SD increase [95% CI 0.211 – 0.277, p<0.001] in PMI and 0.426 SD increase in MI (95% CI 0.395 – 0.457, p<0.001).

**Conclusion:** Altogether, our results suggest that PMI/MI and BMD partially share common biological pathways, and the former may constitute a relevant marker if screening for children with impaired bone health using DPRs.

## INTRODUCTION

Areal bone mineral density (BMD) indicates the mineral content per square centimeter of bone and is used to diagnose osteoporosis and assess fracture risk (1). Genetic and environmental factors such as sex, age, weight, lifestyle factors, fracture history and ancestry have been associated with BMD (2–5). BMD accrual in early life is an important determinant of osteoporosis onset later in life (6,7). The BMD of the mandible is associated with adult features such as dental extraction history, number of teeth present, denture use, shape and thickness of the mandibular cortex (3,8,9). While genetic determinants of mandibular BMD remain unknown, genetic factors already described to influence BMD at other sites (10–16) are likely to contribute to its variation, assuming that a fraction of them will exert systemic effects in general processes such as mineralization. In line with this, previous studies have established a positive correlation between mandibular BMD and BMD at other skeletal sites in the adult population (17).

Dual-energy X-ray Absorptiometry (DXA) is considered the gold standard in the skeletal BMD assessment (18). Likewise, dental panoramic radiographs (DPRs) are widely used in dental practice as a mainstay imaging modality to investigate features of the maxillofacial area. DPRs have been suggested as a useful tool in the early diagnosis of osteoporosis in the elderly, with several mandibular cortical indices developed to assess the quality of mandibular bone. These indices have been associated with the BMD of the peripheral skeleton in the adult population (19–22). In the healthy pediatric population, the relationship between peripheral BMD and mandibular indices remains unknown.

The current study aims to elucidate the genetic architecture of mandibular indices and evaluate the potential relationship between the mandibular indices and total body less-head (TBLH) BMD as determinants of mandibular and skeletal bone health in a multi-ethnic cohort of school-aged children.

## METHODS

### Study population

This study was embedded in the Generation R Study, a population-based, multi-ethnic, prospective cohort study conducted in Rotterdam, the Netherlands. The study focuses on environmental and genomic factors influencing children’s health, growth, and developmental outcomes from early pregnancy until young adulthood (23). Both participants and their parents provided informed consent to participate in this study. The Generation R Study was approved by the Medical Ethical Committee of the Erasmus Medical Centre, Rotterdam, the Netherlands (MEC-2015-749).

### Participant characteristics

Sex and the date of birth of every participant were collected from the medical records and hospital registries. Age was calculated as the difference between the (fourth) phase visit date, and the date of birth. Height and weight were measured by trained research technicians using standardized procedures and used to calculate body mass index (BMI). Whole-body DXA scans (iDXA scanner, GE Healthcare, Madison, WI, USA) were performed according to the manufacturer’s protocols and analyzed using the enCORE software (v13.60). Procedures of quality assurance have been described earlier (24).

### Continentally defined ethnicity of the participants

Participants were divided into three major continentally defined groups: European (Dutch, North African, Turkish, American, other European, Oceanic), African (Antillean, Surinamese-Creole, Cape Verdian and other African), or Asian (Surinamese-Hindustani, Indonesian, other Asian), based on the country of birth of their parents following the classification of Statistics Netherlands (25). This partition was used as individuals from different backgrounds present different BMD levels, as it was established children of different ethnic background have different BMD values as well as risk of fracture (26).

### Puberty

A puberty score was generated based on a pubertal development status questionnaire (27). The questionnaire, which is a revision of a well-known Pubertal Development Scale (28), comprises 5 items (3 general and 2 sex-specific) regarding the growth spurt, growth of new hair on the face or other sites different than the head, skin and/or breast changes, changes in the voice depth and presence of menstruation.

### Mandibular indices

DPRs were performed using the orthopantomograph® OP200D (Oldelft Benelux B.V., Veenendaal, the Netherlands). Scans as well as measurements of mental indices (MI) and panoramic mandibular indices (PMI superior and inferior) were performed using the Viewbox 4 software (dHAL Software, Kifissia, Greece) (29). A detailed description of the measurements used is provided in the supplement. To evaluate the reliability of the MI and PMI measurements, we employed intra-class and inter-class correlations (for a single observer and two observers, respectively), and a paired T-test in a subset of 150 DPRs that were re-evaluated by the same and another trained dentist. The reliability of the PMI superior and MI measurements showed excellent (intra-class correlation coefficient 0.911 [95% CI 0.879 – 0.934], and 0.904 [95% CI 0.871 – 0.930], respectively), and good inter-class correlation coefficient (0.766 [95% CI 0.690 – 0.824], and 0.812 [95% CI 0.750 – 0.861], respectively). Similarly, PMI inferior demonstrated an excellent intra-class correlation coefficient of 0.907 (95% CI 0.872 – 0.932), and a moderate inter-class correlation of 0.728 (95% CI 0.596 – 0.814). The paired T-test did not show significant differences between the two subsets of measurements neither for PMI superior nor MI (both in case of a single observer or two observers) (all p>0.3). Conversely, there were differences between the two sets of measurements for PMI inferior (p_single_observer_=0.013, p_two_observers_<0.001). Therefore, we decided to focus on PMI superior and MI in the current study.

### Genotyping

The Generation R Study was genotyped in two waves. The first and largest subset of participants was genotyped using Human610K and Human 660W genotyping arrays whereas the second was genotyped using the GSA-MD v2 genotyping arrays (Illumina, Inc., San Diego, CA, USA). Phasing, imputation, and quality control steps for the first subset are described in detail elsewhere (30). Information on quality control, phasing, and imputation of the second subset is provided in the supplement. The first 20 genomic principal components were calculated for the joint study population (both subsets). SNPs imputed to the 1000 Genomes Project Phase III Version 5 reference panel exceeding minor allele frequency (MAF) greater or equal to 5% and imputation quality greater or equal to 30% were included in the association analysis. Genetic ancestral background was estimated for children with genotype data using ADMIXTURE (31,32). Children were assigned to one of the ancestral populations based on the highest fraction of the estimated ancestral proportion (>50%). Those where none exceeded the threshold were defined as having admixed ancestral background.

Children allocated to the European ancestral population were included in the polygenic risk score (PGS) analysis to avoid population stratification (33,34).

### Polygenic score analysis

A weighted PGS was calculated per participant as the sum of products of dosages of BMD increasing alleles for 81 independent SNPs, each multiplied by its corresponding effect size as obtained in a large GWAS meta-analysis (10). In the analyses, mandibular cortical indices were compared between the two groups of participants allocated to the extremes of the described BMD PGS distribution. The analysis assumes that confounders across these two groups are randomized (35).

This partition was used as individuals from different backgrounds present different BMD levels, as it was established children of different ethnic background have different BMD values as well as risk of fracture (26).

### Statistical analyses

Baseline characteristics of the participants were compared across sexes using Chi-squared and T-tests. GWAS of mandibular indices were performed separately for each genotyped subset of the Generation R participants using a linear mixed model adjusted for sex, age, and first 10 genomic principal components using RVTESTS (36). Quality control of the association summary statistics was performed with EasyQC (37). Inverse-variance weighted fixed-effect meta-analysis of the two subsets was then performed using METAL (38). The association between BMD and the mandibular indices was tested following two complementary approaches. First, we tested the relationship between standardized BMD and mandibular cortical thickness measurements using linear regression. In a basic model, the associations were adjusted for age, sex, and continental background. Then, associations were additionally adjusted for height and weight. A sensitivity analysis was carried out by adding a pubertal score as a covariate, in the participants where this data was available. Second, we used a PGS approach, using the above explained approach in children from European ancestral background. Briefly, BMD was first regressed on the generated BMD-PGS, and the F-statistic of the association was noted. Then, genotyped participants were stratified into 15% extremes of the PGS distribution. Participants’ characteristics across the bottom and top extremes were compared using a T-test. Differences in mandibular indices between participants of different continental groups were assessed in an unadjusted linear regression model with Europeans as the reference category.

## RESULTS

### Study population

Our observational study included 3,518 children (1,696 boys; 48.2%) with mean age 13.63 (SE 0.38) years, for whom complete data, including TBLH-BMD, mandibular cortical thickness, anthropometric measurements and continentally defined ethnicity measurements were available. In total, 2,879 children were classified as European (81.8%), 430 as African (12.2%), and 209 as Asian (5.9%) based on geographical ancestry.

Boys were on average taller, had thinner mandibular cortical measurements (both PMI and MI), and had lower BMI and TBLH-BMD values as compared to girls (p≤0.001). No significant differences in age, weight, or ancestral percentages were observed across sexes (Table I).

**Table I.**
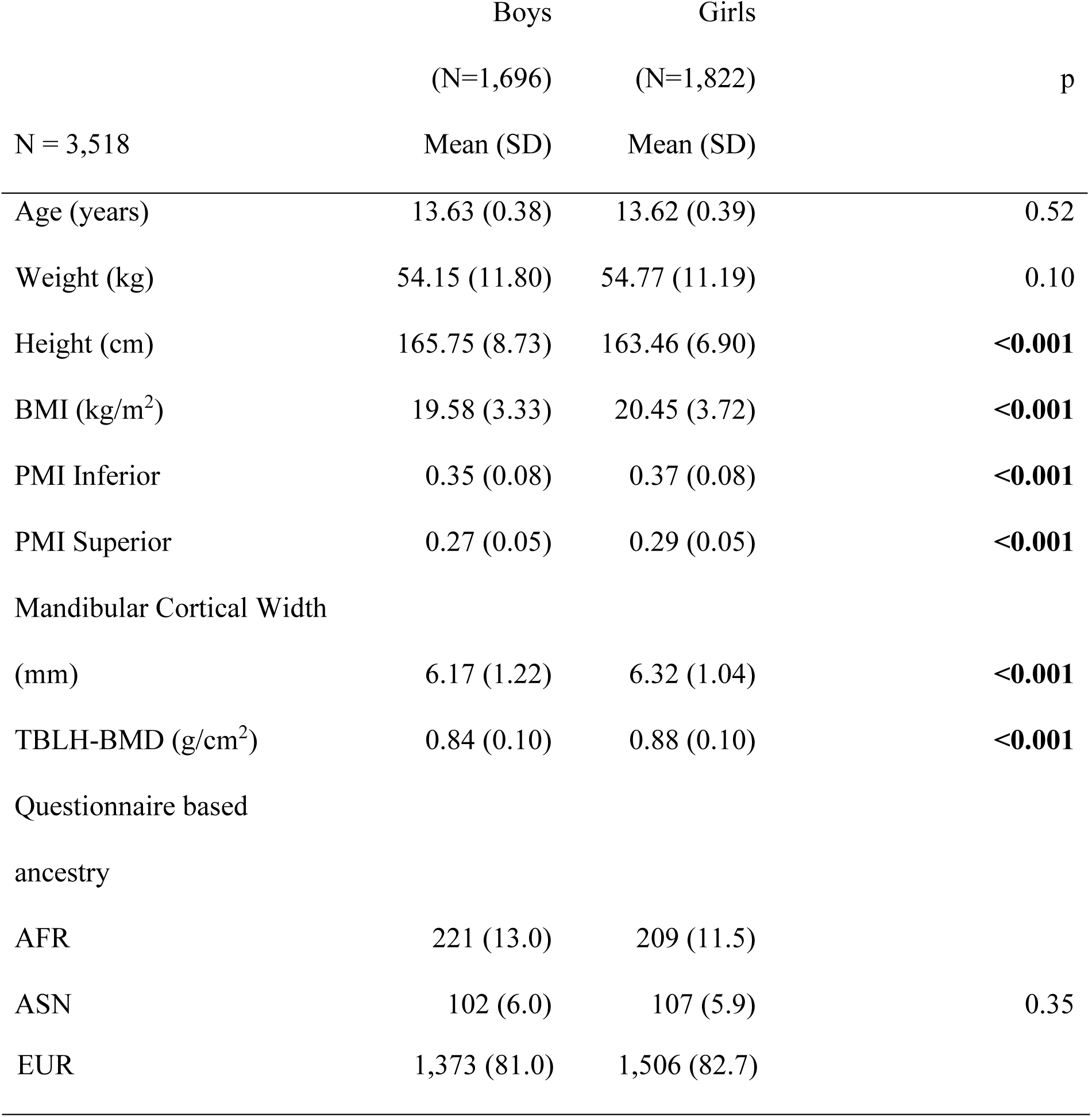
Characteristics of the study population stratified by sex. N – sample size, SD – standard deviation, BMI – body mass index, TBLH-BMD – total body-less head bone mineral density, PMI – panoramic mandibular index, MI – mental index, EUR – European ancestry AFR – African ancestry, ASN – Asian ancestry, *count (percentage), significant p-values are presented in bold

|  | Boys | Girls | p |
| --- | --- | --- | --- |
|  | (N=1,696) | (N=1,822) |  |
| N = 3,518 | Mean (SD) | Mean (SD) |  |
| Age (years) | 13.63 (0.38) | 13.62 (0.39) | 0.52 |
| Weight (kg) | 54.15 (11.80) | 54.77 (11.19) | 0.10 |
| Height (cm) | 165.75 (8.73) | 163.46 (6.90) | **<0.001** |
| BMI (kg/m<sup>2</sup>) | 19.58 (3.33) | 20.45 (3.72) | **<0.001** |
| PMI Inferior | 0.35 (0.08) | 0.37 (0.08) | **<0.001** |
| PMI Superior | 0.27 (0.05) | 0.29 (0.05) | **<0.001** |
| Mandibular Cortical Width |  |  |  |
| (mm) | 6.17 (1.22) | 6.32 (1.04) | **<0.001** |
| TBLH-BMD (g/cm<sup>2</sup>) | 0.84 (0.10) | 0.88 (0.10) | **<0.001** |
| Questionnaire based |  |  |  |
| ancestry |  |  |  |
| AFR | 221 (13.0) | 209 (11.5) |  |
| ASN | 102 (6.0) | 107 (5.9) | 0.35 |

|  |  |  |
| --- | --- | --- |
| EUR | 1,373 (81.0) | 1,506 (82.7) |

### Mandibular indices genome-wide association studies

The PMI-GWAS meta-analysis (N=3,324) identified the *ODF3/BET1L/RIC8A/SIRT3* locus on chromosome 11 (lead SNP rs11604127-T: MAF=0.18, B=0.21, SE=0.03, p=2.53×10^-9^) (Figure I). The phenotypic variance explained by this variant was 1.07%. Also, the MI-GWAS meta-analysis identified this locus with the lead SNP in high linkage disequilibrium (r^2^=0.93) with rs11604127-T (rs55966801-C: B=-0.33, SE=0.04, p=7.30×10^-20^). Results of variants reaching genome-wide significance (p<5×10^-8^) for each respective meta-analysis are provided in the supplement.

**Figure I.**
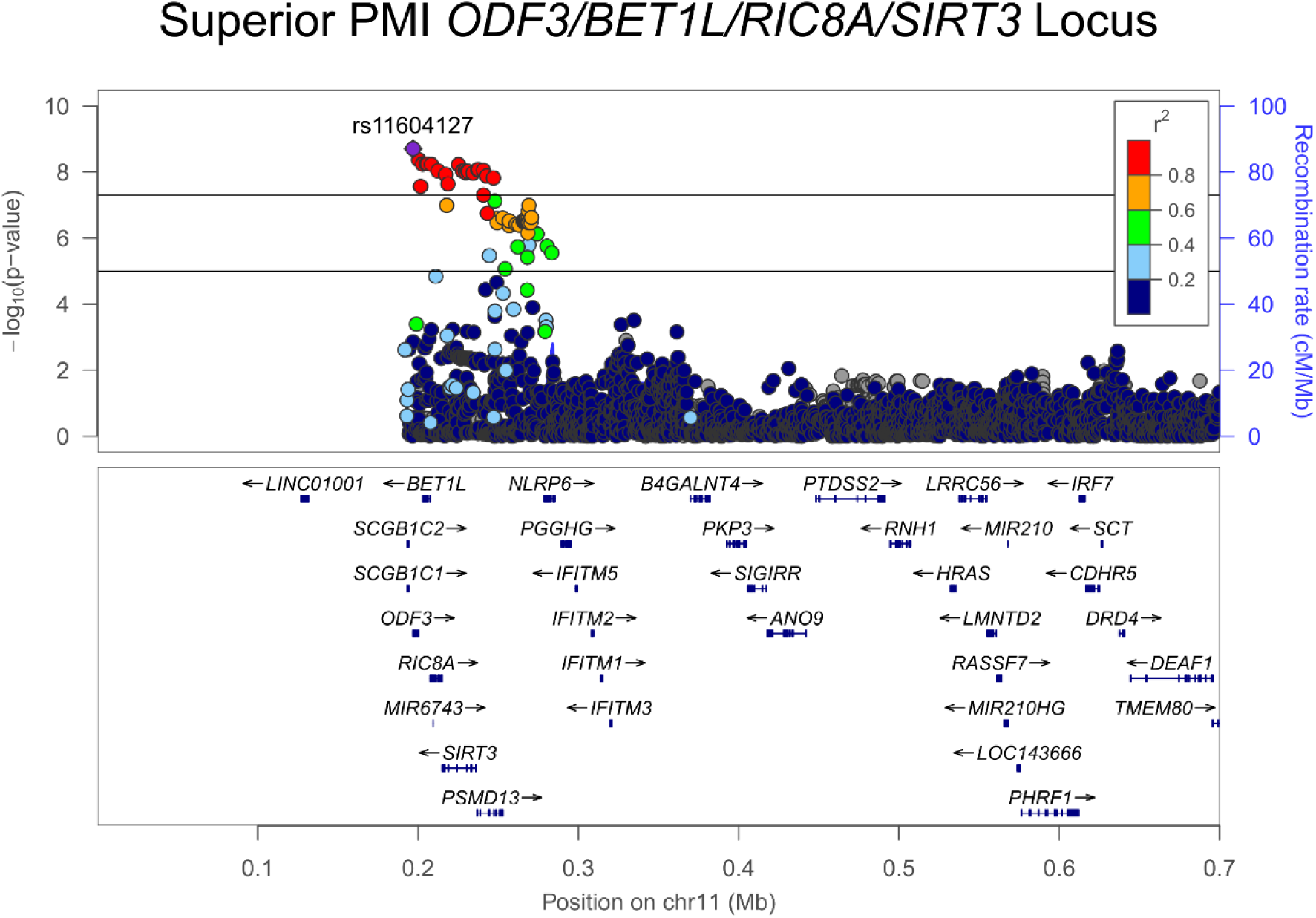
Regional plot for the *ODF3/BET1L/RIC8A/SIRT3* locus associated with PMI superior. The purple diamond is the leading variant (the variant with the lowest p-value). Circles flanking the diamond are color-coded according to their respective linkage disequilibrium measured as a square of correlation to the leading variant. Recombination rates and linkage disequilibrium values are calculated based on the EUR-population (1000 Genomes Project Phase 3). The X-axis represents the genomic position on the 11^th^ chromosome. The y-axis represents the -log10 of the p-values of associations.

### Mandibular indices association with BMD Observational association

After adjusting for age, sex, and geographical ancestry, one TBLH-BMD SD increase was associated with a 0.244 SD increase of PMI superior (95% CI 0.211 – 0.277, p<0.001), and a 0.426 SD increase of MI (95% CI 0.395 – 0.457, p<0.001). Additional adjustments for weight and height did not substantially affect these findings (data not shown), while additional adjustment for puberty attenuated the association (N=2,655, 75.5% of the original sample population). One TBLH-BMD SD was associated with 0.206 SD increase in PMI superior (95% CI 0.150 – 0.260, p<0.001) and a 0.285 SD increase of MI (95% CI 0.233 – 0.337, p<0.001).

### PGS analysis

The weighted TBLH-BMD PGS (N=2,716) was strongly associated with BMD (F-statistic = 80.8). Children in the highest 15% extreme (N=408) of the weighted PGS distribution had significantly higher PMI superior as compared to those in the lowest 15% extreme (mean difference: 0.011 [95% CI 0.005 – 0.019], p<0.001) and MI (mean difference: 0.317 [95% CI 0.167 – 0.467], p<0.001). No differences were observed in sex percentage, age, weight or height (all p>0.1) across the two strata (Table II).

**Table II.** Mandibular cortical indices and other anthropometric measurements presented across the bottom and top 15% of TBLH-BMD weighted PGS. Analysis was performed on participants genetically determined to be European (N=2,716). N – sample size, BMI – body mass index, PMI – panoramic mandibular index, MI – mental index, SD – standard deviation, *count (percentage), significant p-values are presented in bold

|  | Bottom 15% | Top 15% |  |
| --- | --- | --- | --- |
|  | N = 408 | N = 408 |  |
| N=2,716 | Mean (SD) | Mean (SD) | p |
| Sex (Girls)* | 211 (51.7) | 220 (53.9) | 0.58 |
| Age | 13.61 (0.34) | 13.64 (0.37) | 0.25 |
| Weight (kg) | 53.98 (10.51) | 53.82 (11.15) | 0.83 |
| Height (cm) | 164.83 (8.10) | 164.67 (7.32) | 0.77 |
| BMI | 19.80 (3.31) | 19.76 (3.40) | 0.87 |
| PMI Superior | 0.28 (0.05) | 0.29 (0.05) | **0.001** |
| MI (mm) | 6.04 (1.09) | 6.36 (1.09) | **<0.001** |
| PMI Inferior | 0.35 (0.07) | 0.37 (0.08) | **0.002** |

### Ethnic group results

Children classified as having African geographic ethnic background showed thicker mandibular cortices as compared to European children for both PMI superior and MI (0.019 [95% CI 0.014 – 0.024], p<0.001 and 0.494 [95% CI 0.380 – 0.670], p<0.001, respectively). Asian children showed higher MI as compared to Europeans (0.159 [95% CI 0.002 – 0.317], p=0.046).

## DISCUSSION

The present study explored the genetic architecture of the MI and the PMI superior, two mandibular cortical thickness indices in a large multi-ethnic cohort of more than 3000 healthy pubescent children, participants of the Generation R Study. It also established the relation between these indices and BMD in this pediatric cohort, using both an observational and a genetic approach. Our GWAS yielded one genome-wide significant locus mapping to the *ODF3/BET1L*/*RIC8A*/*SIRT3* region. Moreover, our results show a significant association between the mandibular indices and TBLH-BMD even after the adjustment for age, sex, ancestry, height, weight, and puberty. Such an association was also confirmed by leveraging the participant genetic information and knowledge of BMD architecture.

Notwithstanding the small sample size (N=3,3324), our GWAS meta-analyses discovered a PMI/MI locus (*ODF3*/*BET1L*/*RIC8A*/*SIRT3*). Despite differences in the leading SNP for PMI and MI GWAS, these markers were in high LD (r^2^>0.8), suggesting they both underlie a single association signal. This locus has been associated with total body and skull BMD (10,16). Moreover, both variants are associated with changes in the expression of *BET1L*, *SIRT3*, *RIC8A*, *NLRP6* and *SCGB1C1* across multiple tissues (39). Variants mapping to this region may have a systemic effect on bone mineralization. A *SIRT3* deletion attenuated bone loss/resorption caused by estrogen deficiency in mice (40). At the same time, *BET1L* has been repeatedly associated with the increased risk of uterine leiomyoma (41–43), which itself was linked to changes in BMD in women of different ancestries (44,45). Also, shared causal variants were shown to regulate *RIC8A* expression in osteoclasts and play a role in skull bone mineral density (16). Nevertheless, as our GWAS includes discovery and replication populations originating from the same study cohort, future, better-powered efforts should replicate our GWAS findings.

We observed a positive significant association between mandibular cortex thickness and TBLH-BMD in the studied pediatric population. Likewise, over the last decades, numerous studies have shown that the deterioration of the mandibular cortex is associated with bone mineral loss and osteoporosis in the elderly (20,22,46–48). Therefore, DPRs, widely performed in dental clinical practices, have been suggested as a valuable tool in screening patients at risk for osteoporosis (48). To the best of our knowledge, only one previous study investigated PMI as a parameter associated with bone health in children (49). However, this study was carried out in data from 43 HIV-infected children. In line with our findings, decreased mandibular cortex was observed in HIV-positive children with low BMD (51). Nevertheless, no statistical analysis was reported to validate this observation. Using the genetic data of Generation R participants, we ratified the positive relationship between mandibular indexes and BMD. Children in the highest BMD PGS bin had mandibular cortical indices that are on average greater than those in the lowest BMD PGS bin, in contrast to confounding factors randomizing across the two sets of participants. This suggests that TBLH-BMD and mandibular thickness share to a certain extent underlying biology.

We observed higher PMI and MI values in girls as compared to boys. This contrasts with past results from a healthy elderly population showing no significant sex differences in mandibular cortical thickness (50) or mandibular angular cortex (51). However, it aligns with the latest results established in a larger adult population (52). Data describing a sexual dimorphism in mandibular growth and development is scarce. Earlier research showed that boys (N=10) had significantly thicker mandibular cortices than girls (N=14) at the mean age of 13.5 years (not observed in younger age groups) (53), which was later replicated (54). The authors of the former study suggested that pubertal accelerated growth results in greater mandibular height and a thicker mandibular cortex in males. Our results indeed suggest that girls have shorter mandibular height and as a result greater PMI superior.

Children of African ancestral background show a thicker mandibular cortex than children of European ancestral background. This is in line with a previous study (N=353; age range 30-79 years) in which ticker mandibular cortices were described in African individuals (50). On top of this, African children are more advanced in bone and tooth development than their non-African peers (24,32,55) and show higher BMD (24,26). It has been suggested that these differences could correspond to distinct evolutionary challenges that different ancestries were exposed to (31).

Although we established a strong correlation and shared biology between mandibular indices and BMD, utilizing the former is not free of challenges. Scoring mandibular indices on DPRs is currently time-consuming and, as such, unlikely to be implemented in the dental clinical practice despite PMI/MI’s potential diagnostic value. Nonetheless, artificial intelligence (AI) might help circumvent this burden(56). Leite *et al*. have recently introduced a new AI-based tool for accurate and quick tooth detection and segmentation on DPRs (57). This development may also enable a quick and accurate evaluation of the mandibular cortex.

The Generation R Study is, to the best of our knowledge, the only population-based multi-ethnic pediatric cohort having DPRs available. This allows us to investigate DPR-derived traits (e.g., oligo- and hypodontia, dental age, mandibular thickness) in a large setting. Unfortunately, without other cohorts having the same assessments, we cannot replicate our findings at present. Other limitations of our study include the use of DPRs to assess mandibular indices, which are known to be prone to certain distortions. However, this distortion affects to a greater extent the posterior region of DPRs (premolars/molars) as compared to the anterior region (incisors) (58).

In conclusion, this study describes a positive relationship between thinner mandibular cortices and lower BMD in a healthy pediatric population. It additionally suggests a shared biological background between these two traits. Our results suggest that mandibular measurements might be useful to identify children in need of a detailed bone health assessment. An early diagnosis of low BMD would help develop strategies allowing children to reach optimal peak bone mass. This is the first study evaluating these indices in the pediatric population, therefore we favor future replication of our findings.

## Data Availability

All data produced in the present work are contained in the manuscript. Summary statistics of GWAS meta-analysis is made available for download online. http://www.gefos.org/?q=content/mandibular-coritical-indices-gwas-2024

## ACKNOWLEDGEMENTS

We gratefully acknowledge the contribution of participants, hospitals and their staff, and pharmacies in Rotterdam involved in the Generation R Study. Generation and management of GWAS genotype data for the Generation R Study was done at the Genomics Core Facility, Department of Internal Medicine at Erasmus MC. We thank Pascal Arp, Gaby van Dijk, Marijn Verkerk, Samuel Ghatan, Dr. Carolina Medina-Gomez, Dr. Linda Broer and Jard de Vries for their help in creating, managing and QC the GWAS database.

## SUPPLEMENT

### Mandibular indices measurements

DPRs were performed during the same visit as DXA scans, using the orthopantomograph® OP200D (Oldelft Benelux B.V., Veenendaal, the Netherlands). Before scanning, participants were instructed to remove glasses, elastic bands from the hair, and any jewelry that may influence the scan. A chin rest was provided when participants were missing front teeth or had malocclusion. Otherwise, a bite block was used. Scans were then performed following the manufacturer’s specifications. A trained dentist assessed the mandibular cortical thickness, providing bilateral measurements for mental index (MI) and panoramic mandibular indices (PMI superior and inferior) on DPRs. The arithmetic means for each of the three bilateral measurements were established when possible. When only unilateral measurements were available, those from the sinistral side of the mandible were used. MI was measured as the cortical thickness on an axis that crossed the approximate center of the mental foramen and formed a 90° angle with an axis tangential to the lower mandibular rim. The PMI superior was calculated as the ratio of the MI and the distance between the mental foramen’s upper border and the mandible’s lower border, measured on the same vertical axis. The PMI inferior was calculated in the same manner, only using the distance measured to the lower border of the mental foramen (instead of the upper one).

### Genotyping, quality control and imputation of GENR4 dataset

Initial QC consisted of the exclusion of samples with call rate below 97.5%, excess of heterozygosity (more than 4 standard deviations from the mean), sex mismatches and/or genetic duplicates. In addition, variants with call rate below 97.5% and Hardy-Weinberg equilibrium (HWE) deviation (p-value < 1×10^-7^) were excluded. ZCall was used on previously uncalled genotypes aiming to improve the call rate of less-frequent to rare variants (Goldstein et al PMID: 22843986) after which filtering on missingness was more stringent (>99%) for both variants and samples. Lastly, comparison of reported familiar relationships based on questionnaires across both subsets and genetic analyses of familiar relationships resulted in detection and exclusion of sample swaps. The first 20 genomic principal components were calculated for all participants of both datasets simultaneously. Roughly 27,000 independent and common SNPs (minor allele frequency > 0.05) that were in common between GENR3, GENR4, and the 1000 Genomes Project Phase III version 5 (1000 GP; 1000 Genomes Project Consortium PMID:26432245) data were used in the calculation. GENR4 genotypes were phased using ShapeIT v2 r904 (Delaneau PMID:25653097), followed by imputation using minimac4 (Das PMID:27571263) with reference to the 1000 GP.

## Supplementary GWAS results

**Table SI.** Genome-wide significant variants in the PMI-GWAS meta-analysis. All effect sizes (B) are reported for the effect allele (EA). Chr – Chromosome, BP – chromosomal position on genome build 37, OA – Other allele, Freq – Frequency of the effect allele, SE – standard error, P – p-value, N – sample size

| Chr | BP | EA | OA | Freq | B | SE | P | N |
| --- | --- | --- | --- | --- | --- | --- | --- | --- |
| 11 | 196944 | t | c | 0.1768 | 0.2089 | 0.0351 | 2.526e-09 | 3324 |
| 11 | 200343 | t | c | 0.8328 | -0.2129 | 0.0363 | 4.422e-09 | 3324 |
| 11 | 202812 | a | g | 0.1750 | 0.1911 | 0.0334 | 1.051e-08 | 3324 |
| 11 | 205021 | c | g | 0.8249 | -0.1910 | 0.0334 | 1.061e-08 | 3324 |
| 11 | 205889 | t | c | 0.8247 | -0.1909 | 0.0334 | 1.069e-08 | 3324 |
| 11 | 202856 | a | g | 0.1751 | 0.1909 | 0.0334 | 1.083e-08 | 3324 |
| 11 | 208235 | a | g | 0.1753 | 0.1908 | 0.0334 | 1.091e-08 | 3324 |
| 11 | 225180 | a | g | 0.1714 | 0.1951 | 0.0342 | 1.191e-08 | 3324 |
| 11 | 237737 | a | t | 0.1774 | 0.1888 | 0.0334 | 1.621e-08 | 3324 |
| 11 | 237209 | a | g | 0.1774 | 0.1887 | 0.0334 | 1.642e-08 | 3324 |
| 11 | 212408 | t | c | 0.1770 | 0.1883 | 0.0334 | 1.664e-08 | 3324 |
| 11 | 229302 | a | g | 0.1761 | 0.1887 | 0.0334 | 1.683e-08 | 3324 |
| 11 | 240754 | a | c | 0.8230 | -0.1888 | 0.0335 | 1.714e-08 | 3324 |
| 11 | 227798 | a | t | 0.1759 | 0.1885 | 0.0334 | 1.729e-08 | 3324 |
| 11 | 230951 | a | g | 0.1758 | 0.1885 | 0.0335 | 1.776e-08 | 3324 |
| 11 | 231305 | a | g | 0.1759 | 0.1885 | 0.0335 | 1.786e-08 | 3324 |
| 11 | 226083 | d | i | 0.1762 | 0.1882 | 0.0334 | 1.827e-08 | 3324 |
| 11 | 229811 | t | g | 0.8237 | -0.1878 | 0.0335 | 1.983e-08 | 3324 |
| 11 | 234349 | t | c | 0.8245 | -0.1882 | 0.0335 | 2.032e-08 | 3324 |
| 11 | 217171 | t | c | 0.1752 | 0.1879 | 0.0335 | 2.034e-08 | 3324 |
| 11 | 242859 | a | g | 0.8207 | -0.1845 | 0.0330 | 2.306e-08 | 3324 |
| 11 | 247028 | a | g | 0.1804 | 0.1848 | 0.0332 | 2.503e-08 | 3324 |
| 11 | 218640 | a | g | 0.2188 | 0.1730 | 0.0316 | 4.45e-08 | 3324 |

**Table SII.** Genome-wide significant variants in the MI-GWAS meta-analysis. All effect sizes (B) are reported for the effect allele (EA). Chr – Chromosome, BP – chromosomal position on genome build 37, OA – Other allele, Freq – Frequency of the effect allele, SE – standard error, P – p-value, N – sample size

| Chr | BP | EA | OA | Freq | B | SE | P | N |
| --- | --- | --- | --- | --- | --- | --- | --- | --- |
| 11 | 200343 | t | c | 0.8328 | -0.3277 | 0.0359 | 7.298e-20 | 3324 |
| 11 | 196944 | t | c | 0.1768 | 0.3153 | 0.0347 | 1.034e-19 | 3324 |
| 11 | 205021 | c | g | 0.8249 | -0.2982 | 0.0330 | 1.792e-19 | 3324 |
| 11 | 205889 | t | c | 0.8248 | -0.2982 | 0.0330 | 1.792e-19 | 3324 |
| 11 | 208235 | a | g | 0.1753 | 0.2983 | 0.0331 | 1.82e-19 | 3324 |
| 11 | 202812 | a | g | 0.1749 | 0.2983 | 0.0331 | 1.862e-19 | 3324 |
| 11 | 202856 | a | g | 0.1750 | 0.2982 | 0.0331 | 1.889e-19 | 3324 |
| 11 | 212408 | t | c | 0.1770 | 0.2962 | 0.0330 | 2.999e-19 | 3324 |
| 11 | 227798 | a | t | 0.1758 | 0.2959 | 0.0331 | 4.086e-19 | 3324 |
| 11 | 217171 | t | c | 0.1751 | 0.2963 | 0.0332 | 4.13e-19 | 3324 |
| 11 | 229302 | a | g | 0.1760 | 0.2958 | 0.0331 | 4.213e-19 | 3324 |
| 11 | 230951 | a | g | 0.1757 | 0.2957 | 0.0331 | 4.657e-19 | 3324 |
| 11 | 226083 | d | i | 0.1761 | 0.2954 | 0.0331 | 4.733e-19 | 3324 |
| 11 | 231305 | a | g | 0.1758 | 0.2955 | 0.0331 | 4.927e-19 | 3324 |
| 11 | 225180 | a | g | 0.1713 | 0.3017 | 0.0339 | 5.293e-19 | 3324 |
| 11 | 229811 | t | g | 0.8238 | -0.2947 | 0.0331 | 5.779e-19 | 3324 |
| 11 | 234349 | t | c | 0.8247 | -0.2950 | 0.0332 | 6.629e-19 | 3324 |
| 11 | 269129 | t | c | 0.2059 | 0.2765 | 0.0311 | 6.811e-19 | 3324 |
| 11 | 237737 | a | t | 0.1773 | 0.2909 | 0.0331 | 1.52e-18 | 3324 |
| 11 | 237209 | a | g | 0.1773 | 0.2908 | 0.0331 | 1.551e-18 | 3324 |
| 11 | 201584 | a | t | 0.1628 | 0.3134 | 0.0357 | 1.581e-18 | 3324 |
| 11 | 247028 | a | g | 0.1803 | 0.2881 | 0.0328 | 1.718e-18 | 3324 |
| 11 | 240754 | a | c | 0.8232 | -0.2909 | 0.0332 | 1.744e-18 | 3324 |
| 11 | 242859 | a | g | 0.8208 | -0.2859 | 0.0327 | 2.237e-18 | 3324 |
| 11 | 270330 | a | t | 0.8042 | -0.2804 | 0.0321 | 2.564e-18 | 3324 |
| 11 | 268927 | t | c | 0.2004 | 0.2756 | 0.0316 | 2.934e-18 | 3324 |
| 11 | 268940 | t | c | 0.8001 | -0.2750 | 0.0316 | 3.352e-18 | 3324 |
| 11 | 261209 | t | c | 0.1979 | 0.2731 | 0.0316 | 4.992e-18 | 3324 |
| 11 | 262930 | t | c | 0.1981 | 0.2729 | 0.0315 | 5.193e-18 | 3324 |
| 11 | 252851 | a | g | 0.8027 | -0.2722 | 0.0315 | 5.25e-18 | 3324 |
| 11 | 256978 | t | c | 0.1978 | 0.2719 | 0.0315 | 5.562e-18 | 3324 |
| 11 | 270715 | t | c | 0.7957 | -0.2686 | 0.0312 | 8.025e-18 | 3324 |
| 11 | 274181 | a | c | 0.1892 | 0.2843 | 0.0331 | 8.068e-18 | 3324 |
| 11 | 268718 | a | g | 0.7773 | -0.2574 | 0.0303 | 1.853e-17 | 3324 |
| 11 | 280464 | c | g | 0.8123 | -0.2910 | 0.0344 | 2.534e-17 | 3324 |
| 11 | 247905 | a | c | 0.1666 | 0.3439 | 0.0409 | 4.454e-17 | 3324 |
| 11 | 217814 | t | g | 0.1600 | 0.3003 | 0.0358 | 4.759e-17 | 3324 |
| 11 | 267568 | a | g | 0.2220 | 0.2542 | 0.0304 | 6.022e-17 | 3324 |
| 11 | 218640 | a | g | 0.2187 | 0.2610 | 0.0313 | 7.719e-17 | 3324 |
| 11 | 268628 | a | g | 0.2222 | 0.2528 | 0.0303 | 7.752e-17 | 3324 |
| 11 | 249408 | a | g | 0.1849 | 0.2675 | 0.0324 | 1.416e-16 | 3324 |
| 11 | 249131 | a | g | 0.1841 | 0.2678 | 0.0324 | 1.55e-16 | 3324 |
| 11 | 256976 | a | c | 0.2161 | 0.2502 | 0.0304 | 1.97e-16 | 3324 |
| 11 | 267250 | a | g | 0.7540 | -0.2451 | 0.0298 | 2.045e-16 | 3324 |
| 11 | 266957 | t | g | 0.2460 | 0.2448 | 0.0298 | 2.175e-16 | 3324 |
| 11 | 266921 | a | g | 0.7541 | -0.2447 | 0.0298 | 2.218e-16 | 3324 |
| 11 | 266804 | t | c | 0.2459 | 0.2446 | 0.0298 | 2.269e-16 | 3324 |
| 11 | 268430 | c | g | 0.7740 | -0.2465 | 0.0301 | 2.594e-16 | 3324 |
| 11 | 265777 | t | g | 0.2456 | 0.2439 | 0.0298 | 2.661e-16 | 3324 |
| 11 | 266248 | t | c | 0.7541 | -0.2439 | 0.0298 | 2.67e-16 | 3324 |
| 11 | 266315 | t | c | 0.7544 | -0.2438 | 0.0298 | 2.915e-16 | 3324 |
| 11 | 265602 | a | g | 0.7542 | -0.2431 | 0.0298 | 3.213e-16 | 3324 |
| 11 | 263868 | d | i | 0.2455 | 0.2417 | 0.0297 | 4.395e-16 | 3324 |
| 11 | 218055 | d | i | 0.9460 | -0.7129 | 0.0884 | 7.212e-16 | 3324 |
| 11 | 240844 | t | c | 0.7818 | -0.2439 | 0.0310 | 3.31e-15 | 3324 |
| 11 | 283415 | a | g | 0.1669 | 0.2939 | 0.0376 | 5.456e-15 | 3324 |
| 11 | 268183 | t | c | 0.2019 | 0.2646 | 0.0339 | 6.391e-15 | 3324 |
| 11 | 243268 | t | g | 0.7786 | -0.2356 | 0.0304 | 9.623e-15 | 3324 |
| 11 | 242126 | d | i | 0.2114 | 0.2454 | 0.0318 | 1.133e-14 | 3324 |
| 11 | 268020 | a | g | 0.7242 | -0.2167 | 0.0293 | 1.427e-13 | 3324 |
| 11 | 262191 | a | c | 0.6829 | -0.2049 | 0.0284 | 5.234e-13 | 3324 |
| 11 | 254421 | a | g | 0.7082 | -0.1923 | 0.0279 | 5.121e-12 | 3324 |
| 11 | 279214 | c | g | 0.2744 | 0.2011 | 0.0305 | 4.409e-11 | 3324 |
| 11 | 270071 | t | c | 0.5258 | -0.1560 | 0.0252 | 5.759e-10 | 3324 |
| 11 | 269147 | t | c | 0.5097 | -0.1513 | 0.0248 | 1.09e-09 | 3324 |
| 11 | 244552 | a | g | 0.4336 | 0.1557 | 0.0257 | 1.4e-09 | 3324 |
| 11 | 279798 | a | g | 0.3496 | 0.2175 | 0.0361 | 1.71e-09 | 2370 |
| 11 | 259575 | t | c | 0.0887 | 0.2916 | 0.0484 | 1.724e-09 | 3324 |
| 11 | 249105 | a | g | 0.0489 | 0.4545 | 0.0761 | 2.303e-09 | 3324 |
| 11 | 280000 | a | g | 0.3469 | 0.2138 | 0.0363 | 3.92e-09 | 2370 |
| 11 | 198986 | t | c | 0.5499 | -0.1484 | 0.0256 | 6.365e-09 | 3324 |
| 11 | 247998 | t | c | 0.1047 | 0.2614 | 0.0454 | 8.427e-09 | 3324 |
| 11 | 253072 | t | c | 0.5744 | -0.1452 | 0.0254 | 1.111e-08 | 3324 |
| 11 | 247906 | c | g | 0.5371 | 0.1853 | 0.0332 | 2.455e-08 | 3324 |
| 11 | 211015 | t | c | 0.4149 | -0.1401 | 0.0257 | 4.871e-08 | 3324 |

## Abbreviations

PMI: Panoramic mandibular index
MI: Mental index
DPR: dental panoramic radiograph
TBLH-BMD: total body-less head bone mineral density
DXA: dual-energy X-ray absorptiometry
GWAS: genome-wide association study
PGS: polygenic score
SD: standard deviation

